# A Breath-Based In Vitro Diagnostics for Lower Respiratory Tract Infection

**DOI:** 10.1101/2023.09.18.23295728

**Authors:** Dapeng Chen, Marek A. Mirski, Emily R. Caton, Kiana M. Kiser, Caroline R. Haddaway, Maximilian S. Cetta, Shuo Chen, Wayne A. Bryden, Michael McLoughlin

## Abstract

Lower Respiratory Tract Infections (LRTIs) represent the leading cause of death due to infectious diseases. Current diagnostic modalities primarily depend on clinical symptoms and lack specificity, especially in light of common colonization without overt infection. To address this, we developed a noninvasive diagnostic approach that employs BreathBiomics™, an advanced human breath sampling system, to detect protease activities induced by bacterial infection in the lower respiratory tract. Specifically, we engineered a high-sensitivity and high-specificity molecular sensor for human neutrophil elastase (HNE). The sensor undergoes cleavage in the presence of HNE, an event that is subsequently detected via Matrix-Assisted Laser Desorption/Ionization Time of Flight Mass Spectrometry (MALDI-TOF MS). Application of this methodology to clinical samples, breath specimens collected from intubated patients with LRTIs, demonstrated the detection of the cleaved sensor by MALDI-TOF MS. Our findings indicate that this novel approach offers a noninvasive and specific diagnostic strategy for people with LRTIs.

**Significance**
The potential for using human breath for noninvasive disease detection and diagnosis has long been recognized, yet the lack of effective biomolecular sampling technologies has hindered progress. To address this limitation, we developed BreathBiomics™, an advanced sampling system designed to efficiently capture biomolecules in human exhaled breath. By focusing on protease dysregulation, an established event induced by bacterial infections, we demonstrated that BreathBiomics™ can capture proteases and facilitate their subsequent activity-based detection for the diagnosis of LRTI. We verified the assay’s sensitivity and clinical applicability through empirical studies. Our work marks a significant advancement by providing the first viable pathway for the development of in vitro diagnostic assays leveraging human breath for disease detection and diagnosis.

## Introduction

Early treatment with short-course antibiotics is crucial for managing lower respiratory tract infections (LRTIs) in mechanically ventilated patients. Currently, diagnostic approaches relying on clinical criteria lack specificity (1). Molecular diagnostics like multiplex (polymerase chain reaction) PCR assays have been developed but cannot distinguish between bacterial colonization and infection (2). To address this, host-response markers have been investigated in bronchoalveolar lavage fluid (BALF) samples, which offer higher specificity but are invasive and thus unsuitable for routine testing (3). Therefore, there is a critical need for non-invasive diagnostic methods for LRTIs in mechanically ventilated patients.

The balance between proteases and anti-proteases is crucial for maintaining airway homeostasis, particularly in bacterial colonization in the lower respiratory tract (4). Multiple studies have shown that an imbalanced immune response can lead to the excessive activation of alveolar macrophages and neutrophils, releasing proteases like matrix metalloproteinases (MMPs) and human neutrophil elastase (HNE) (3-5). Therefore, detecting protease activity can be used as a diagnostic strategy for LRTI (6).

Previously, we developed a sampling technology using a novel capture mechanism, BreathBiomics™, designed to efficiently capture biomolecules like proteins from exhaled human breath (7). In the current study, we elaborated on this by developing an in vitro assay focused on detecting HNE activity within human breath, utilizing an ultra-sensitive sensor specific to HNE. Taken together, our findings indicate that this method holds substantial promise for an in vitro diagnostic approach for LRTIs.

## Results

### HNE Sensor Design and Diagnosis Strategy

The proposed HNE sensor comprises a polymer linker, an amino acid substrate to HNE, and a cleavable small molecule (Figure 1A). An HNE sensor was designed to follow this concept. The polymer linker is a polyethylene glycol (PEG) 36. As previously reported, the HNE substrate includes unnatural amino acids, which show sensitivity and specificity to HNE (8). The small molecule that can be cleaved is 7-Amino-4-carbamoylmethylcoumarin (ACC) (Figure 1B). The diagnostic methodology involves four critical steps (Figure 1C). Initially, HNE is induced during bacterial infections, and is present in the biofluids of the lower respiratory tract. It is emitted through airway reopening mechanisms into human exhaled breath. Utilizing BreathBiomics™ technology, the HNE can be captured in the human exhaled breath. Subsequently, the captured HNE is extracted and subjected to *in-vitro* incubation with the specially designed HNE sensor. In the presence of HNE, the sensor undergoes a cleavage reaction that can be identified via Matrix-Assisted Laser Desorption/Ionization-Time of Flight Mass Spectrometry (MALDI-TOF MS). Theoretically, the MALDI-TOF spectra should show two mass peaks corresponding to the intact sensor and its cleavage product, respectively.

**Figure 1.**
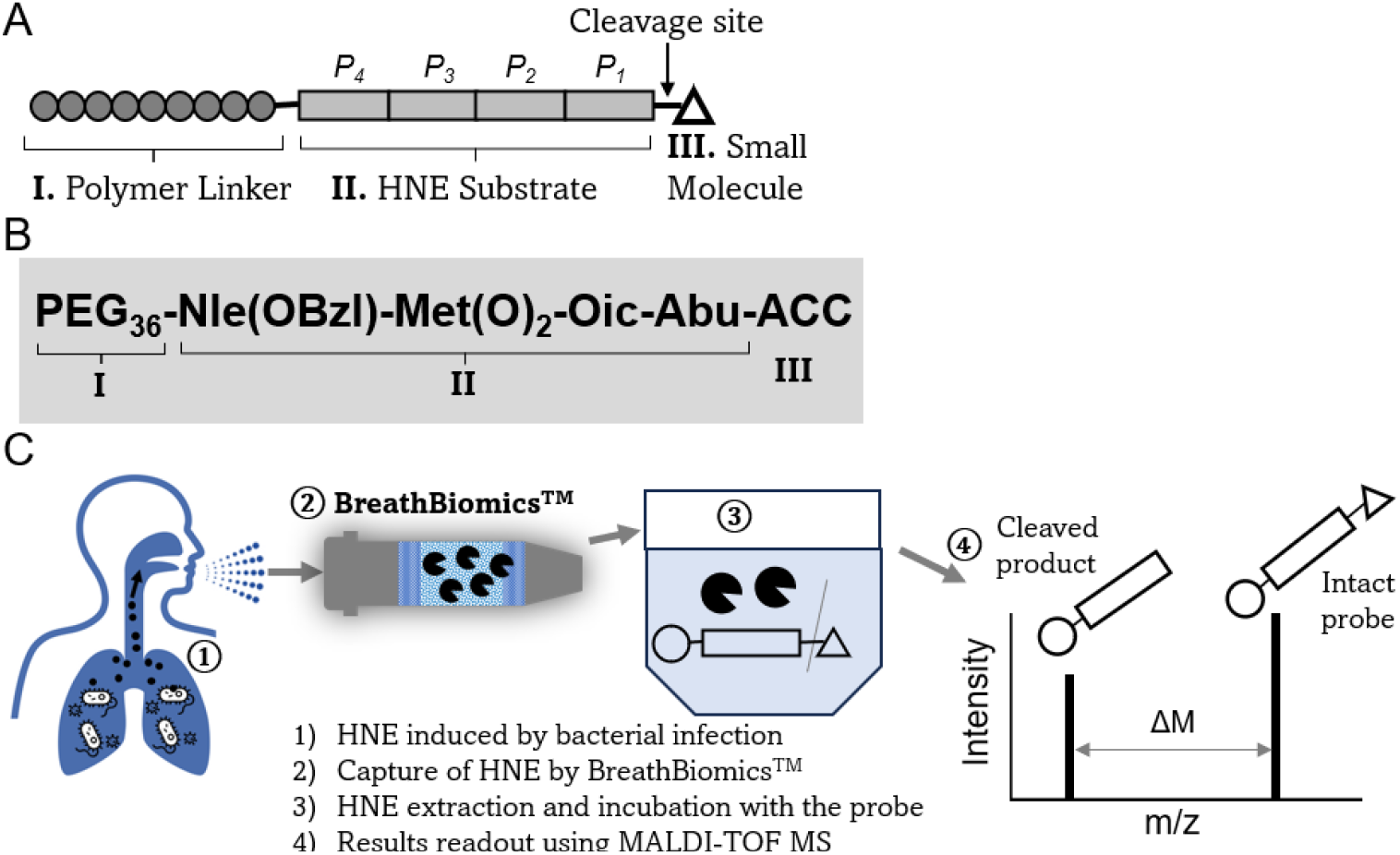
Schematic of the approach and overview. (A) The concept for designing the HNE sensor. (B) The customized HNE sensor used for the in vitro diagnostics strategy in this study. (C) Overview of the in vitro diagnostics strategy for LRTI.

### Testing the HNE Sensor Using Recombinant Mouse NE

Our study evaluated the customized HNE sensor using a recombinant mouse ELA2 protein. A concentration of 100 μM of the HNE sensor was incubated with 50 ng of ELA2 protein at 37°C at various time intervals, followed by analysis via MALDI-TOF MS. Beginning at the 10-minute mark of incubation, additional peaks, besides the intact sensor mass peak of 2557.6 m/z, with m/z ratios of 2357.6 and 2379.5 were observed in the MALDI-TOF mass spectra (dark and red stars, Figure 2A). The identities of the intact sensor and the cleavage products were further corroborated using high-resolution mass spectrometry (Figure 2B). Given that the reaction buffer contained sodium ions, mass peaks indicative of sodium adducts were also detected. Notably, the peak corresponding to the intact sensor disappeared after 180 minutes of incubation, suggesting complete cleavage into the reaction products. Our findings reveal a time-dependent relationship, as the ratio of the intact sensor to its cleavage products showed a progressive increase throughout the incubation period (Figure 2C).

**Figure 2.**
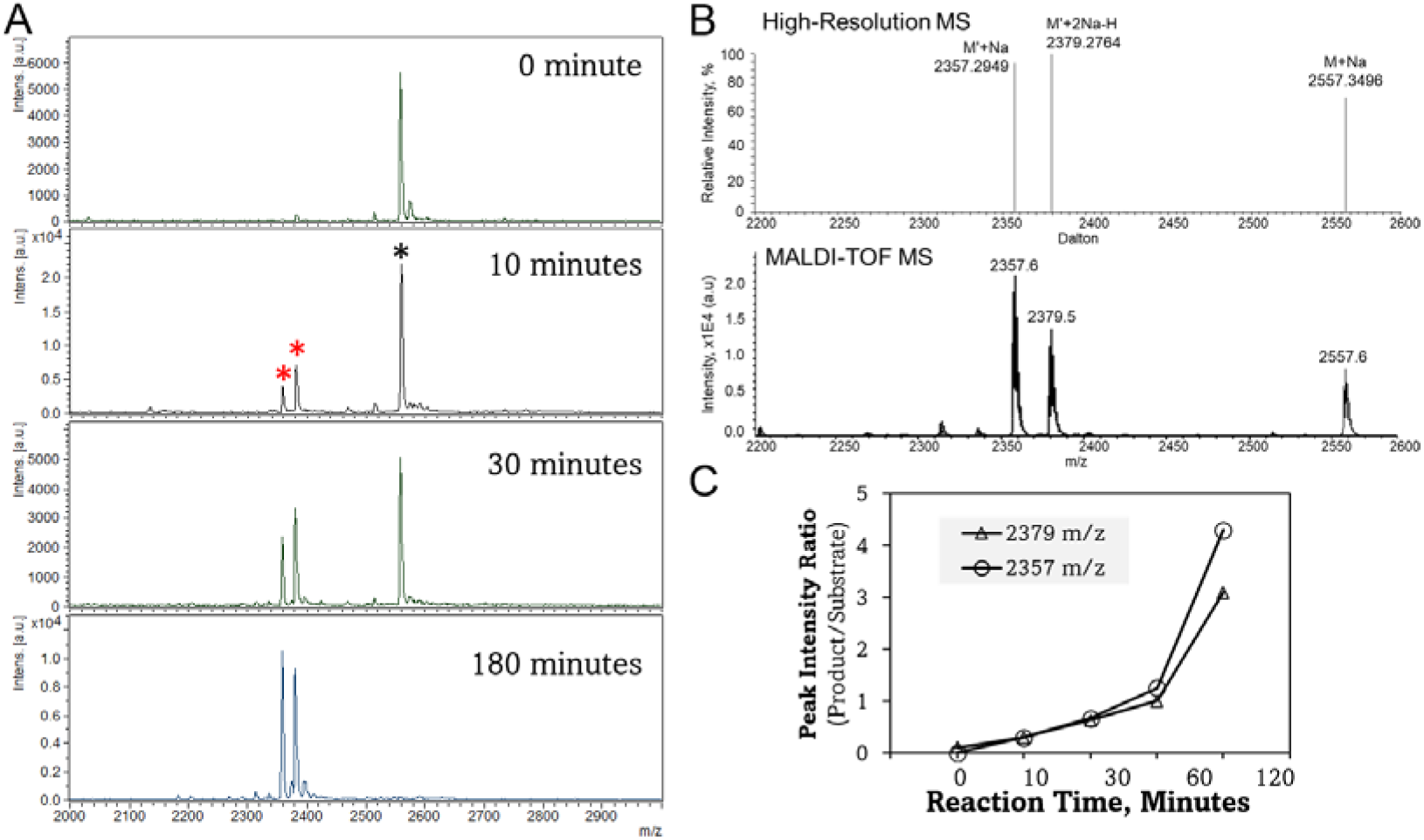
Detection of the customized HNE sensor and the cleavage products using MALDI-TOF MS. (A) MALDI-TOF mass spectra of the HNE sensor and the cleavage products in different reaction time points. (B) Characterization of the HNE sensor and the cleavage products by high-resolution mass spectrometry. (C) Ratio of the cleavage products to the HNE sensor during the reaction.

### Detection of The Cleavage Products of HNE in Exhaled Breath Samples from People with Mechanical Ventilation

We extended our approach to analyze human exhaled breath samples, which were collected from participants situated in the Intensive Care Unit (ICU) at Johns Hopkins Hospital (JHH) (Figure 3A). Our findings indicate that the levels of cleavage products were elevated in participants diagnosed with LRTI, suggesting our approach using breath and the customized HNE sensor could serve as a promising diagnostic tool for monitoring and detecting LRTI in critical care settings (dark and red stars, Figure 3B).

**Figure 3.**
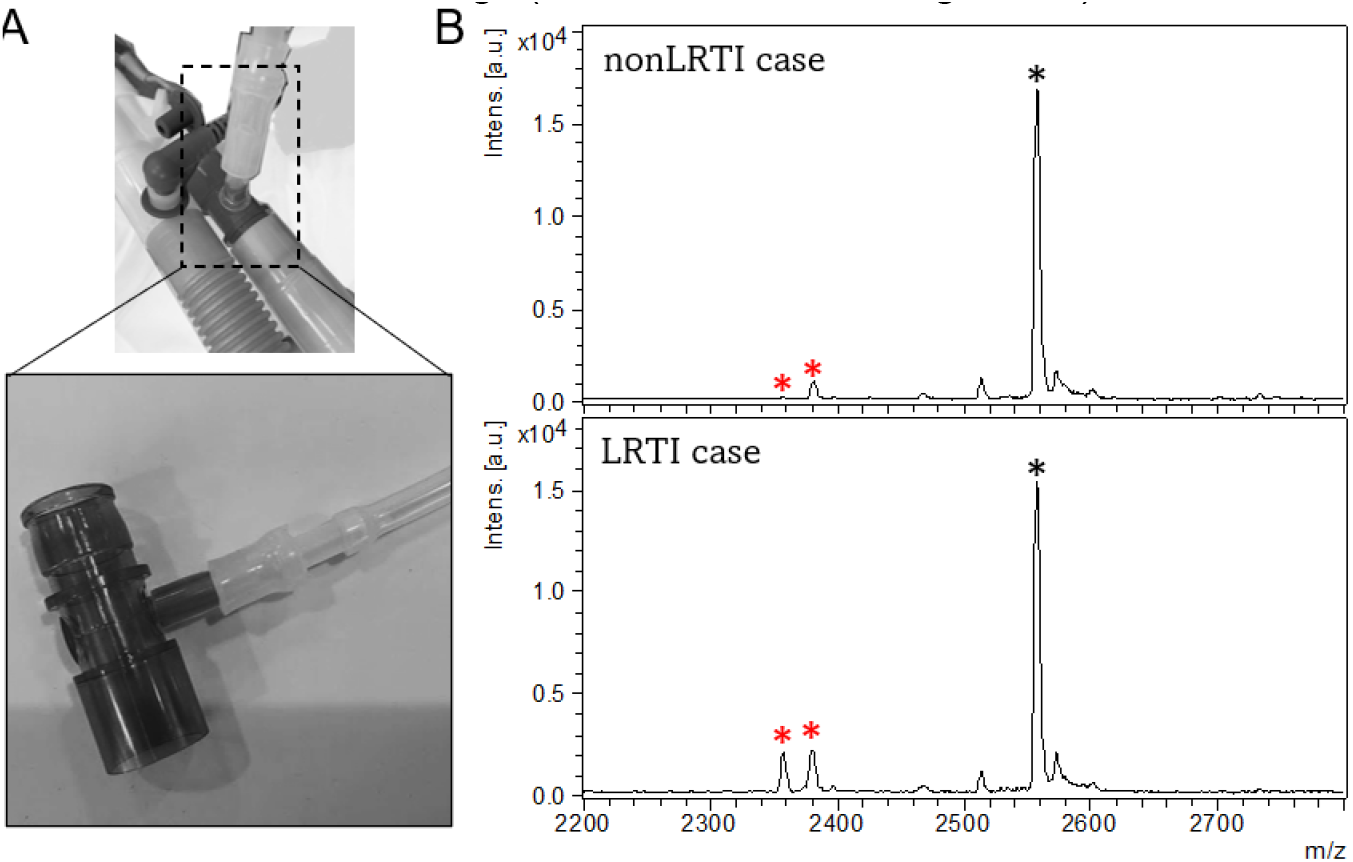
Application of our approach to clinical samples. (A) Human exhaled breath collection at the ICU at JHH using BreathBiomics™. (B) MALTI-TOF mass spectra of HNE sensor and cleavage products in nonLRTI and LRTI cases.

## Discussion

Previous studies demonstrated the dysregulation of neutrophil elastase (HNE) in severe respiratory conditions, particularly in community-acquired pneumonia (CAP) and ventilator-associated pneumonia (VAP) (3, 9,10). Van Eeden et al. (1991) observed a transient functional inactivation of α1-proteinase inhibitor (α1PI) in ICU-treated CAP patients, which was not detected in patients with milder forms of CAP. In addition, this α1PI dysfunction normalized within four weeks post-discharge for survivors, implicating it in pulmonary tissue damage and adverse clinical outcomes (9). Greene et al. (2003) identified elevated HNE concentrations in the infected lung lobes of 17 unilateral CAP patients. While levels of antiproteases such as Alpha-1 antitrypsin (A1AT) and secretory leucoprotease inhibitor (SLPI) also rose, their enzymatic inhibitory activities were suboptimal, pointing to a disruption in the protease-antiprotease equilibrium (10). Wilkinson et al. (2012) observed significant elevations of HNE, MMP-8, and MMP-9 in bronchoalveolar lavage (BAL) fluid from VAP cases compared to non-VAP and control subjects (3). These studies highlight the pivotal role of protease dysregulation in the pathophysiology of severe respiratory infections and suggest that HNE can be used as a promising biomarker with high sensitivity and specificity for the diagnosis of LRTI.

Chan et al. (2020) demonstrated the use of engineered nanoparticles as breath biomarkers for detecting respiratory diseases, specifically focusing on HNE (5). These nanosensors release volatile reporters upon cleavage by HNE. When administered intrapulmonary into mouse models with acute lung inflammation, these reporters are expelled in the breath and detectable via mass spectrometry within as little as 10 minutes post-administration. The study demonstrated the high sensitivity and practicality of this approach, which uses the non-invasive and easy nature of breath sampling to allow real-time monitoring of HNE activity for disease diagnosis.

In the current study, we have demonstrated an *in-vitro* diagnostic (IVD) strategy that uses the established activity of HNE induced by bacterial infections. Distinguishing our methodology is the capacity for quantitative analysis: both the unaltered sensor and its cleavage products are detectable. This enables accurate quantification of HNE activity and associated protein levels, offering a unique advantage of the diagnostic approach.

## Materials and Methods

### Development of the HNE Sensor

The customized HNE sensor was synthesized by CPC Scientific (Sunnyvale, CA).

### HNE Sensor Assay Using Mouse Recombinant NE

Recombinant mouse ELA2 protein was purchased from R&D Systems (Catalog #: 4517-SE). Two buffers were used: an Activation Buffer composed of 50 mM MES and 50 mM NaCl at pH 5.5, and an Assay Buffer containing 50 mM Tris, 1 M NaCl, and 0.05% (w/v) Brij-35 at pH 7.5. Recombinant mouse active Cathepsin C/DPPI (rmCathepsin C), purchased from R&D Systems (Catalog # 2336-CY) was used to active recombinant mouse ELA2. The rmELA2 was diluted to a concentration of 50 μg/mL in Activation Buffer, which also contained 50 μg/mL of rmCathepsin C. This solution was incubated for 2 hours at 37 °C to activate the rmELA2. Following activation, the rmELA2 was further diluted to 1 ng/μL using Assay Buffer. The HNE sensor was diluted to 200 μM in Assay Buffer. For the assay, 50 μL of the 1 ng/μL rmELA2 was used. The reaction was initiated by adding 50 μL of the 200 μM substrate at 37°C.

### Characterization of the NE Sensor and the Cleavage Products Using MALDI-TOF MS

Mass spectrometry analysis was conducted using a Bruker microflex® LRF Mass Spectrometer, operating in positive linear mode. For MALDI plate preparation, 1 μL of the sample was spotted onto the target and allowed to dry. Subsequently, 1 μL of α-cyano-4-hydroxycinnamic acid matrix, prepared at a concentration of 9 mg/mL in 70% acetonitrile, was added on top of the dried sample spot. Once this layer dried, the prepared MALDI plate was inserted into the MALDI-TOF mass spectrometer for analysis. Spectral data was acquired over a mass range of 1000-3000 m/z. An average of 500 laser shots were collected for each spectrum.

### Characterization of the NE Sensor and the Cleavage Products Using high-resolution mass spectrometry

Mass spectrometry analysis was conducted using a Orbitrap Fusion™ Lumos™ Tribrid™ Mass Spectrometer (Thermo Fisher Scientific). Samples were introduced to the ion source by direct infusion. For the data acquisition was conducted in the positive ion mode with a resolution of 60,000. The collected raw data files were deconvoluted and visualized using FreeStyle™ 1.8 software (Thermo Fisher Scientific).

### Exhaled Breath Sample Collection from Study Participant with Mechanical Ventilation

The method for collecting exhaled breath samples from people with mechanical ventilation in the ICU has been described in our previous study (11). The study received approval from the Institutional Review Boards at The Johns Hopkins University School of Medicine (Application Number: IRB00249449), and all study participants provided informed consent. All experimental procedures were executed in accordance with these approved protocols. To collect exhaled air, the BreathBiomics™ column was connected to a tee fitting, installed in the exhaust tubing of the mechanical ventilator, as depicted in Figure 3A. The BreathBiomics™ column was linked to a CO_2_ sensor (Gas Sensing Solutions Ltd, Cumbernauld, UK) and a mini diaphragm pump (Parker Hannifin Corporation, Cleveland, OH). The pump’s flow rate was calibrated to 0.5 liters per minute, and the CO_2_ sensor was utilized to monitor individual levels of exhaled CO_2_ within the exhaust tubing of the mechanical ventilator. Upon completion of sample collection, the columns were disengaged from the ventilator and underwent decontamination procedures in the clinical laboratory at The Johns Hopkins Hospital. The decontaminated columns were subsequently sent to Zeteo Tech’s laboratory for further sample preparation.

### Application of *in vitro* Assay to Human Exhaled Breath Samples

Proteins captured in the collection columns were eluted by adding 300 μL of 70% acetonitrile. Organic solvent was then removed from the samples through an overnight lyophilization process. The resultant dried samples were subsequently reconstituted in 50 μL of high-performance liquid chromatography (HPLC)-grade water. For the bioassay, a 5 μL aliquot of the reconstituted sample was incubated with an equal volume of reaction buffer. This mixture was then combined with 10 μL of the 100 μM HNE sensor. The final reaction assembly was incubated at a temperature of 37°C for a duration of 24 hours.

## Data Availability

All data produced in the present study are available upon reasonable request to the authors.

## Acknowledgments

This work was supported by the National Institute of Allergy and Infectious Diseases (NIAID) of the NIH under Awards R44AI177245.

## Notes

### Competing Interest Statement

D.C., K.M.K, C.R.H., E.R.C., M.S.C., W.A.B., and M.M. have competing interests. D.C., K.M.K., C.R.H., M.S.C., and E.R.C. are employed by Zeteo Tech, Inc. W.A.B. and M.M. serve as chief executive officer and chief technology officer for Zeteo Tech, Inc., respectively. M.A.M. and S.C. have no competing interests. An unpublished U.S. Provisional Patent Application assigned to Zeteo Tech, Inc. was applied based on this research.

### Author Declarations

The study received approval from the Institutional Review Boards at The Johns Hopkins University School of Medicine (Application Number: IRB00249449), and all study participants provided informed consent. All experimental procedures were executed in accordance with these approved protocols.

